# Acute psychosis and the risk of motor vehicle crash

**DOI:** 10.1101/2024.12.10.24318820

**Authors:** John A Staples, Daniel Daly-Grafstein, Mayesha Khan, Shannon Erdelyi, Herbert Chan, William G. Honer, Jeffrey R Brubacher

**Affiliations:** Department of Medicine, University of British Columbia, Vancouver, Canada; Centre for Clinical Epidemiology & Evaluation, Vancouver, Canada; Department of Statistics, University of British Columbia, Vancouver, Canada; Department of Emergency Medicine, University of British Columbia, Vancouver, Canada; Department of Psychiatry, University of British Columbia, Vancouver, Canada; British Columbia Mental Health and Substance Use Services Research Institute, Vancouver, Canada

**Keywords:** Psychotic disorders [MeSH], Psychosis [MeSH], Schizophrenia [MeSH], Traffic crashes, Automobile driving [MeSH], Injury, Trauma, Responsibility analysis, Culpability analysis

## Abstract

**Importance:** Limited empirical evidence guides fitness-to-drive decision-making following an episode of acute psychosis.

**Objective:** To evaluate the association between acute psychosis and subsequent crash risk.

**Design:** Retrospective observational analyses using 20 years of population-based administrative health and driving data. We first assessed the association between psychosis and collisions using a case-crossover design, which controls for relatively fixed individual characteristics like personality, driving experience and routine driving habits. Next, we conducted a responsibility analysis which accounts for the changes in road exposure (miles of driving per month) that might occur after recent hospitalization.

**Setting:** British Columbia, Canada.

**Participants:** Drivers with a police-attended motor vehicle crash, 2000-2016.

**Exposure:** A hospital stay for acute psychosis ending in the 6-week interval prior to crash.

**Main Outcomes and Measures:** The case-crossover analysis examined crash involvement as a driver. The responsibility analysis examined driver responsibility for contributing to their crash. We used logistic regression with adjustment for potential confounders to evaluate associations between outcomes and recent acute psychosis.

**Results:** Among 9842 crashes in the case-crossover analysis, a hospital stay for acute psychosis ended in 199 pre-crash intervals and in 147 control intervals, suggesting acute psychosis was temporally associated with subsequent crash (2.0% vs 1.5% of intervals; adjusted odds ratio (aOR), 1.32; 95%CI, 1.05-1.66; p=0.02). Among 819,348 drivers with a police-attended crash and determinate crash responsibility, 178 of 235 drivers with a recent hospitalization for acute psychosis and 440,543 of 819,113 drivers without recent psychosis were deemed responsible for their crash (75.7% vs 53.8%; aOR, 2.38; 95%CI, 1.75-3.24; p<0.001).

**Conclusions:** The 6-week interval following a hospitalization for acute psychosis is associated with increased odds of crash and increased likelihood of a driver being deemed responsible for contributing to their crash. More stringent temporary driving restrictions after an episode of acute psychosis might reduce crash risk.

**KEY POINTS:** *Questions:* Does a recent episode of acute psychosis increase a driver’s likelihood of being involved in a motor vehicle crash?

*Findings:* Using population-based administrative health and driving data, investigators found that the odds of crash were higher in the first 6 weeks after a hospital stay for acute psychosis than during control periods. A responsibility analysis accounting for changes in road exposure found drivers were also more likely to be deemed responsible for contributing to their crash during this period.

*Meaning:* More stringent driving restrictions in the first 6 weeks after an episode of acute psychosis might reduce crash risk.

## Introduction

Acute psychosis is a disorienting mental state that often requires hospitalization and typically improves after several weeks of psychiatric treatment.^1,2^ Psychosis can impair the perception and judgment required to safely operate a motor vehicle, but extremely limited prior evidence specifically examines this issue. An uncontrolled case series of 94 drivers with psychotic disorder who were killed in vehicle collisions in Finland (1990-2011) found that half had a hospitalization for psychosis in the 3 months prior to death.^3^ We recently published a population-based study found that 2551 crash-involved drivers with schizophrenia were modestly more likely than 805,881 crash-involved drivers without schizophrenia to be deemed responsible for their crash (66.2% vs 53.7%, respectively; aOR, 1.67; 95%CI, 1.53-1.82), but the effect of acute psychosis on crash risk and crash responsibility was not specifically examined.^4^ Most other prior studies of mental illness and crash risk enrolled few patients,^5,6,7^ are several decades old,^5,6,7,8^ focus on a heterogenous mix of chronic mental illnesses,^5,8,9^ and fail to examine acute psychosis as a distinct risk factor for crash.^5,6,7,8,9,10^ As a result, vanishingly little recent empirical evidence informs clinical fitness-to-drive decision-making for patients recovering from an episode of acute psychosis.^10,11,12,3^

Acute psychosis typically occurs among individuals with chronic conditions including schizophrenia, mood disorders, and substance use disorders that can independently influence driving safety.^4^ Individuals experiencing acute psychosis may also substantially increase or decrease their road exposure (the hours or miles of driving per month).^12,13^ Cohort studies on this topic are highly likely to produce biased or misleading results because 1) available data may not capture fundamental baseline differences between drivers with psychosis and control drivers without psychosis, and 2) temporary changes in road exposure during convalescence from acute psychosis might produce a ‘*crash risk per year*’ that obscures a clinically meaningful change in *‘crash risk while driving’*.^14,15^

Other study designs avoid these shortcomings. Case-crossover studies use self-matching to compare an individual’s exposure status in the period immediately before a crash to their exposure status in earlier control periods, thereby avoiding bias arising from the selection of control patients and eliminating confounding from relatively fixed individual characteristics such as genetics, personality, occupation, driving experience, and daily travel routines (**Supplemental Appendix, eFigure 1**).^16,17,18,19,20^ Responsibility analyses use police-reported crash data to categorize crash-involved drivers as cases (deemed ‘responsible’ for their crash) or controls (deemed ‘non-responsible’ for their crash), comparing the prevalence of a potential risk factor between these groups (**eFigure 1**).^21,22,23,24^ Because all individuals are driving when they crash, responsibility analyses inherently account for between-group differences in road exposure.^25,18^

**Figure 1:**
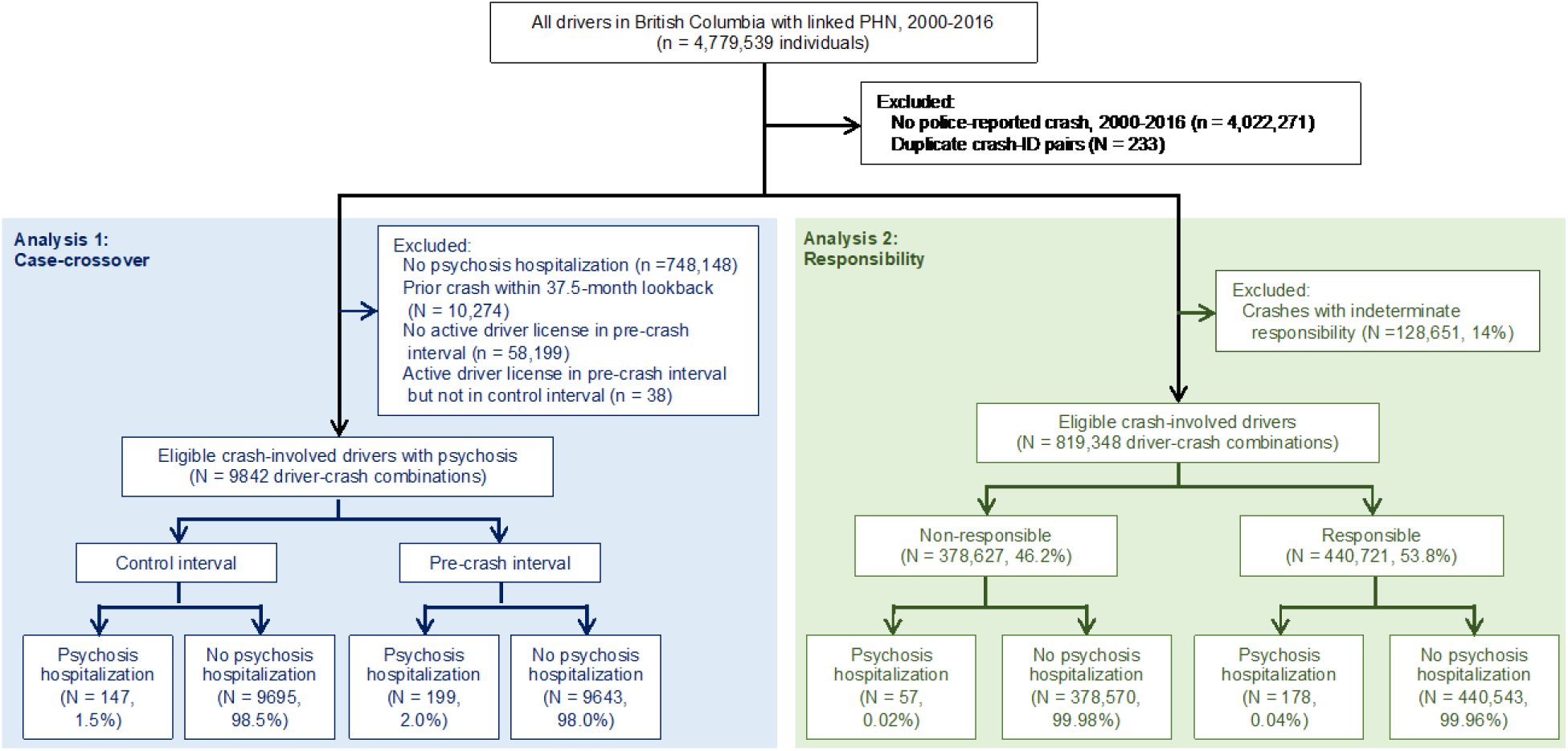
Flow diagram **Legend.** PHN = Personal Health Number; n = individuals; N = unique driver-crash combinations.

Reasoning that evidence of heightened crash risks might prompt psychiatrists to advise temporary cessation of driving, we examined the hypothesis that acute psychosis produces a transient elevation in crash risk that lasts up to 6 weeks after the final day of a psychosis-related hospitalization.

## Methods

### Data sources

We set our study in British Columbia (BC), Canada (2016 population: 4.6 million).^26^ De-identified individual-level data were obtained from population-based administrative health and driving databases (**eMethods 1**).^4,15,17,22,25,27^ We obtained data for all police-attended crashes in BC from the Traffic Accident System (TAS). Police in BC are required to attend all fatal crashes, most serious injury crashes, and some crashes with property damage only. The attending officer completes a structured crash report that identifies each crash-involved driver and records detailed information on the circumstances of the crash. We obtained driver data including license type and traffic contravention history. We used hospitalization and physician billing data to identify medical comorbidities and recent health services use. Medication use was characterized with data on dispensation date and days supplied for all outpatient prescriptions filled at any community pharmacy in BC.

### Study population

The study population included all individuals involved as a driver in a police-attended crash that occurred in BC between 1 January 2000 and 31 December 2016. We excluded individuals if they did not hold a BC driver license or if driving data could not be linked to heath data. By anchoring on involvement as a driver in a police-attended crash, all study analyses pragmatically focus on the clinically-relevant group of individuals who are active drivers at risk of crash. Each anchoring eligible police-attended crash is hereafter termed an ‘index crash’.

### Exposure: Recent episode of acute psychosis

The exposure of interest for both analyses was a recent episode of acute psychosis, defined as a hospitalization for acute psychosis with a discharge date in the 6 week interval immediately prior to the index crash. Hospitalizations with a primary (“most responsible”) diagnosis of acute psychosis were identified using *International Statistical Classification of Diseases and Related Health Problems, Tenth Revision, Canada (ICD-10-CA)* diagnostic codes (**eTable 1**).^28^ We focused on the first 6 weeks after hospital discharge because individuals may continue to experience residual distortions in perception and judgement after discharge, and because hospitalization itself presents a clear opportunity for the provision of driving advice.^3,25,15^

### Analysis 1: Case-crossover analysis

The cohort for the case-crossover analysis included all crash-involved drivers from the study population who had ≥1 police-attended crash as a driver between 1 April 2000 and 31 December 2016 plus ≥1 hospitalization for acute psychosis between 1 January 1997 and 31 December 2016 (**eFigure 1**). The outcome of interest was police-attended crash. For each index crash, we used the crash date (t_0_) to establish a seasonally-matched ‘no crash’ control date exactly 52 weeks earlier (t_0-52wk_). Exposure lookback windows were the 6-week periods before the crash date (‘pre-crash interval’) and the control date (‘control interval’). Case-crossover designs require control intervals to be free of the outcome, so we prespecified that crashes were only eligible if there was no additional crash in the 37.5 months prior to the index crash (to accommodate the main analysis and a sensitivity analysis with a t_0-3y_ control date in the same cohort). Drivers were required to have an active license in all pre-crash and control intervals.

### Drivers could contribute multiple eligible crashes to the analysis

We used conditional logistic regression to compare the likelihood of psychosis hospitalization (exposure; 1=yes, 0=no) in the pre-crash interval to the likelihood of psychosis hospitalization in the control interval, as for a conventional case-control study. We adjusted for potential time-varying confounders: residential neighbourhood income quintile, residential urbanicity, and number of active prescription medications at crash and control dates; and number of impairment-related traffic contraventions, number of non-impairment-related traffic contraventions, number of hospital admissions, and number of physician visits or hospitalizations for ‘alcohol use’ and ‘other substance use’ in a 90-day lookback interval from each crash and control date (**eFigure 2, eTable 2**).

**Figure 2.**
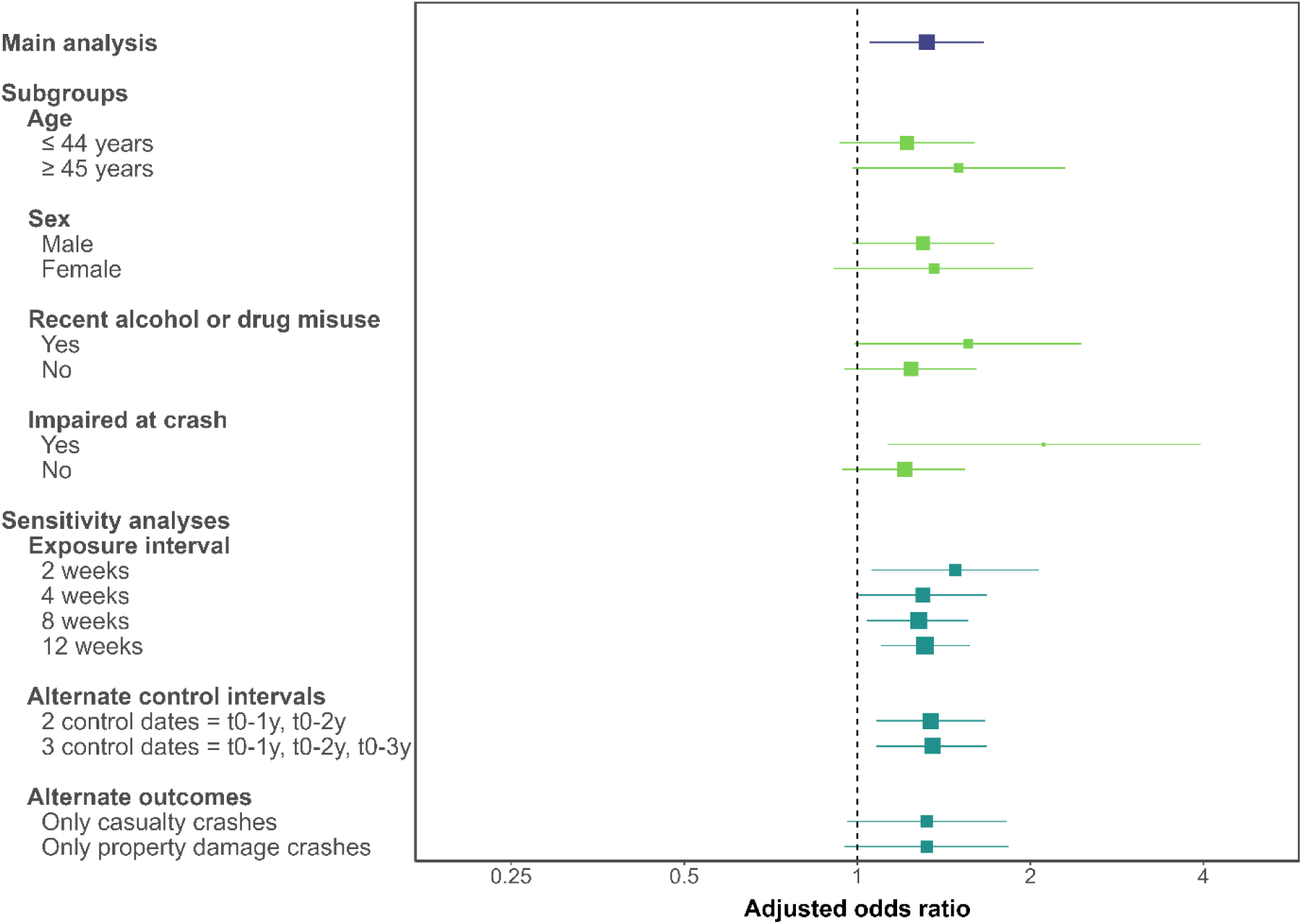
Forest plot of main and subgroup analyses for the case-crossover study. **Legend.** Forest plot of adjusted odds ratios evaluating the association between psychosis and crash for key subgroup and sensitivity analyses. X-axis depicts the adjusted odds ratio on the log scale; squares, the point estimate, with size reflecting the inverse of the standard error; horizontal lines, the 95% confidence interval. Casualty crashes are those that resulted in an injury or fatality. The results of subgroup and sensitivity analyses are generally consistent with results of the main case-crossover analysis.

### Analysis 2: Responsibility analysis

The cohort for the responsibility analysis included all crash-involved drivers from the study population who had ≥1 police-attended crash as a driver between 1 January 2000 and 31 December 2016. The outcome of interest was driver responsibility for the crash (**eFigure 1**). We categorized crash-involved drivers as ‘responsible’ or ‘non-responsible’ for their crash using a validated crash responsibility scoring tool that considers unsafe driving action by the index driver plus six external factors that potentially contribute to a crash: road type, driving conditions, vehicle condition, contribution from other parties, type of collision, and task involved.^21^ When several external factors contribute to a crash (score ≥16), the responsibility tool concludes that the driver could not have reasonably avoided the crash and the driver is deemed ‘non-responsible’ for the crash. Unsafe driving by the index driver or absence of external contributing factors (score ≤13) suggest that the driver should have been able to avoid the crash; the driver is deemed ‘responsible’ for contributing to the crash. Drivers with ‘indeterminate responsibility’ (score 14 or 15) are excluded from further analysis. In this context, crash responsibility is independent of determinations of criminal, civil, or insurance-based responsibility for the crash.

We used logistic regression to examine the association between crash responsibility (outcome; 1=’responsible’, 0=’non-responsible’) and a recent hospitalization for acute psychosis (exposure; 1=yes, 0=no). Our final model adjusted for potential confounders and sources of variability: driver age group and sex; characteristics of the driver’s residential neighborhood including urbanicity, median household income quintile, and regional health authority; hospitalizations or physician visits for substance use in a 3-year lookback; Charlson Comorbidity Index ≥2; driver license type (none, learner, novice, or full); number of years with a full license; number of crashes in a 3-year lookback; number of impairment-related traffic contraventions in a 3-year lookback; number of non-impairment-related traffic contraventions in a 3-year lookback; crash severity (fatality vs injury vs property damage only), location (highway vs rural road vs city streets), and timing (night vs day); documented impairment by alcohol or drugs at the time of crash (yes vs no); and season and calendar year of crash **(eFigure 2, eTable 2)**.^22^

### Additional analyses

We performed exploratory subgroup and sensitivity analyses for both study designs. We also calculated the ‘absolute annual crash rate’ and the ‘responsible crash rate’ (proportion of drivers responsible for their crash) for crash-involved drivers with and without a hospitalization for psychosis.

### Research process

The University of British Columbia Clinical Research Ethics Board approved the study and waived the requirement for individual consent (H12-02678). We did not involve patients or the public in the design, conduct, reporting, or dissemination of our research. Data were deidentified before release to investigators. All inferences, opinions, and conclusions drawn are those of the authors and do not reflect the opinions or policies of the Data Stewards.

## Results

### Analysis 1: Case-crossover analysis

The case-crossover cohort included 9880 crashes among drivers with ≥1 hospitalization for acute psychosis (**Figure 1**). Two thirds of the cohort was male; median age was 38 years (**Table 1**). Almost half of the crashes resulted in an injury, 60 (0.6%) resulted in a fatality, and almost two-thirds involved multiple vehicles, highlighting that crashes often have serious consequences for other road users (**eTable 3**). A total of 38 crashes were dropped from further analysis because the driver lacked an active driver license at either crash or control date.

**Table 1:** Cohort characteristics for the case-crossover analysis. **Legend.** Characteristics of 9880 drivers with at least one hospitalization for acute psychosis during the study interval. “Characteristics that are plausibly relatively fixed between crash and control dates” are presented at the crash date for descriptive purposes only. Drivers who lacked an active license at crash or control date were dropped from further analysis, leaving 9842 crash-involved drivers in the main case-crossover analysis (Figure 1). Characteristics of drivers’ residential neighbourhoods were obtained using census data. p-values compare characteristics between the crash and control dates. *Defined as ≥1 hospitalization or ≥2 physician visits for the condition over the prior 3 years.

| Characteristics | Characteristics at crash date ($t_0$),<br>N= 9880,<br>count (%) | Characteristics at control date ($t_{0-1y}$),<br>N=9880,<br>count (%) | p-value |
| --- | --- | --- | --- |
| **Characteristics that are plausibly relatively fixed between crash and control dates** |  |  |  |
| **Age** |  |  |  |
| . ≤20 years | 909 (9.2) |  |  |
| . 21 to 44 years | 5466 (55.3) |  |  |
| . 45 to 64 years | 2803 (28.4) |  |  |
| . ≥65 years | 702 (7.1) |  |  |
| **Male sex** | 6391 (64.7) |  |  |
| **Medical encounters in prior 3 years for specific conditions\*** |  |  |  |
| . Any psychiatric disorder | 3743 (37.9) |  |  |
| . Problematic drug use | 1082 (11.0) |  |  |
| . Problematic alcohol use | 546 (5.5) |  |  |
| . Hypertension | 450 (4.6) |  |  |
| . Diabetes | 270 (2.7) |  |  |
| **Active driver license at any point in prior 3 years** | 9880 (100) |  |  |
| **Driver license type at time of crash** |  |  |  |
| . Learner | 84 (0.9) |  |  |
| . Novice | 1539 (15.6) |  |  |
| . Full | 8257 (83.6) |  |  |
| **Median years with full license** | 13.8 |  |  |
| **Police-attended crash in prior 3 years** | 1464 (14.8) |  |  |
| **Any traffic contravention in prior 3 years** | 5656 (57.2) |  |  |
| . Speed | 3168 (32.1) |  |  |
| . Alcohol | 1203 (12.2) |  |  |
| . Distraction | 187 (1.9) |  |  |
| **Characteristics that plausibly vary between crash and control dates** |  |  |  |
| **Residential neighborhood household income quintile** |  |  | 0.25 |
| . 1 (lowest income) | 2479 (25.1) | 2434 (24.6) |  |
| . 2 | 2154 (21.8) | 2101 (21.3) |  |
| . 3 | 1815 (18.4) | 1790 (18.1) |  |
| . 4 | 1709 (17.3) | 1757 (17.8) |  |
| . 5 (highest income) | 1434 (14.5) | 1455 (14.7) |  |
| . Missing | 289 (2.9) | 343 (3.5) |  |
| **Residential neighborhood location** |  |  | 0.006 |
| . Urban | 5082 (51.4) | 5053 (51.1) |  |
| . Rural | 4721 (47.8) | 4705 (47.6) |  |
| . Missing | 77 (0.8) | 122 (1.2) |  |
| **Medical encounters in prior 90 days** |  |  |  |
| . ≥1 hospitalization | 679 (6.9) | 574 (5.8) | 0.002 |
| . ≥1 encounter for alcohol use | 26 (0.3) | 25 (0.3) | 1.00 |
| . ≥1 encounter for drug use | 27 (0.3) | 15 (0.2) | 0.09 |
| **Prescription medication fills in prior 90 days** |  |  | <0.001 |
| . 0 or 1 | 4044 (40.9) | 4291 (43.4) |  |
| . ≥2 | 5836 (59.1) | 5589 (56.6) |  |
| **Traffic contraventions in prior 90 days** |  |  |  |
| . Impairment related contravention | 350 (3.5) | 132 (1.3) | <0.001 |
| . Non-impairment related contravention | 2175 (22.0) | 1351 (13.7) | <0.001 |

In the remaining 9842 crashes, discharge from an acute psychosis hospitalization occurred in 199 pre-crash intervals and in 147 control intervals, suggesting that acute psychosis was associated with a significant increase in subsequent crash risk (2.0% vs 1.5% of intervals; adjusted odds ratio [aOR], 1.32, 95%CI, 1.05-1.66, p=0.02; **Figure 2, eTable 4**). Results from subgroup and sensitivity analyses were consistent with the main results (**eTable 5**). The point estimate for the association between crash and recent hospitalization for psychosis was slightly higher with shorter exposure interval lengths, potentially supporting a causal relationship between resolving acute psychosis and crash risk (i.e., for a 2 week lookback, aOR 1.48, 95%CI 1.06-2.07, p=0.02).

### Analysis 2: Responsibility analysis

Among the 947,999 drivers involved in a police-attended crash eligible for the responsibility analysis, 440,721 (46.5%) drivers were deemed responsible and 378,627 (39.9%) were deemed non-responsible for their crash; 128,651 (13.6%) had indeterminate responsibility and were excluded from further analysis (**Figure 1**). Crash-involved drivers had a median age of 39 years and 58% were male (**Table 2**).

**Table 2:**
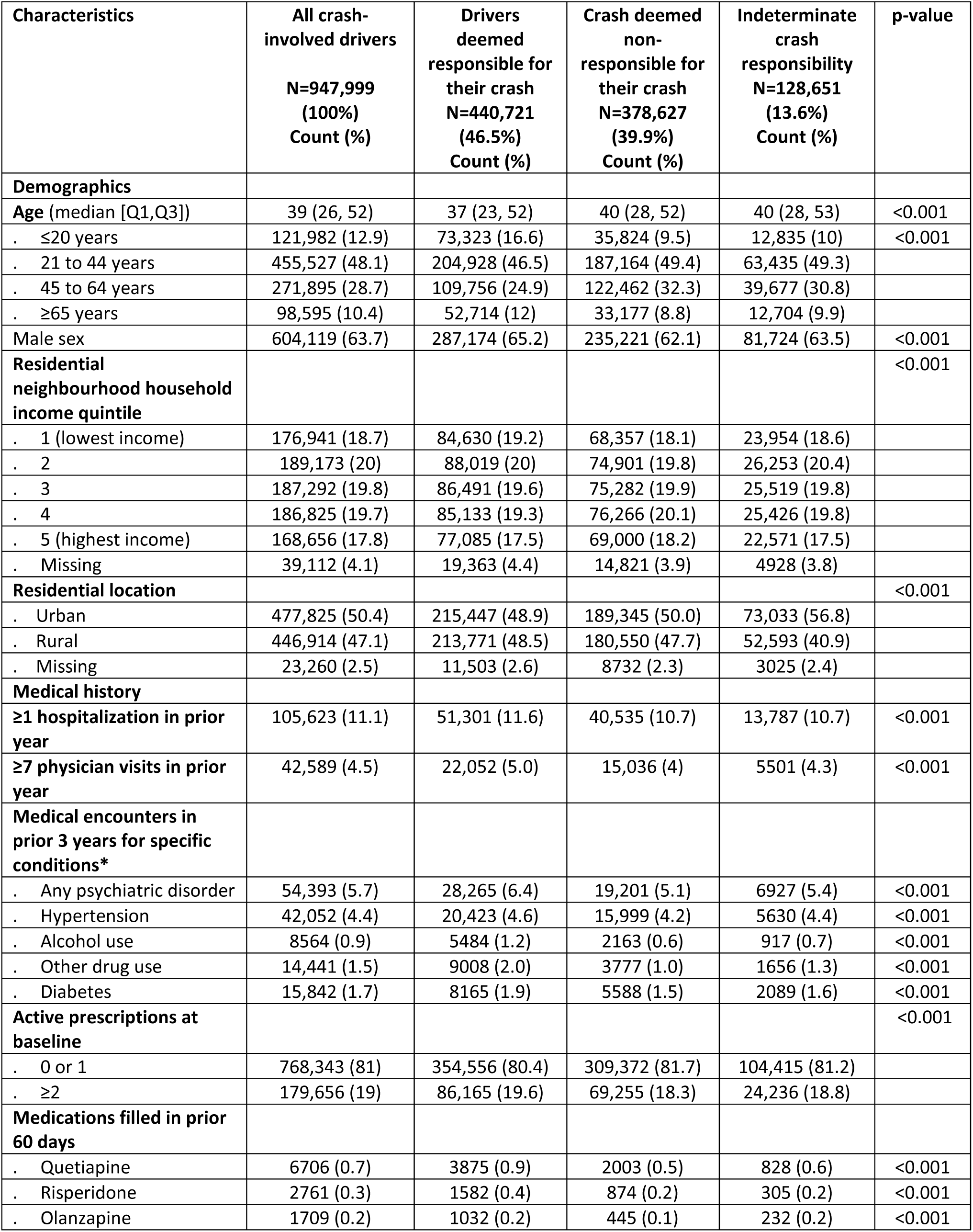
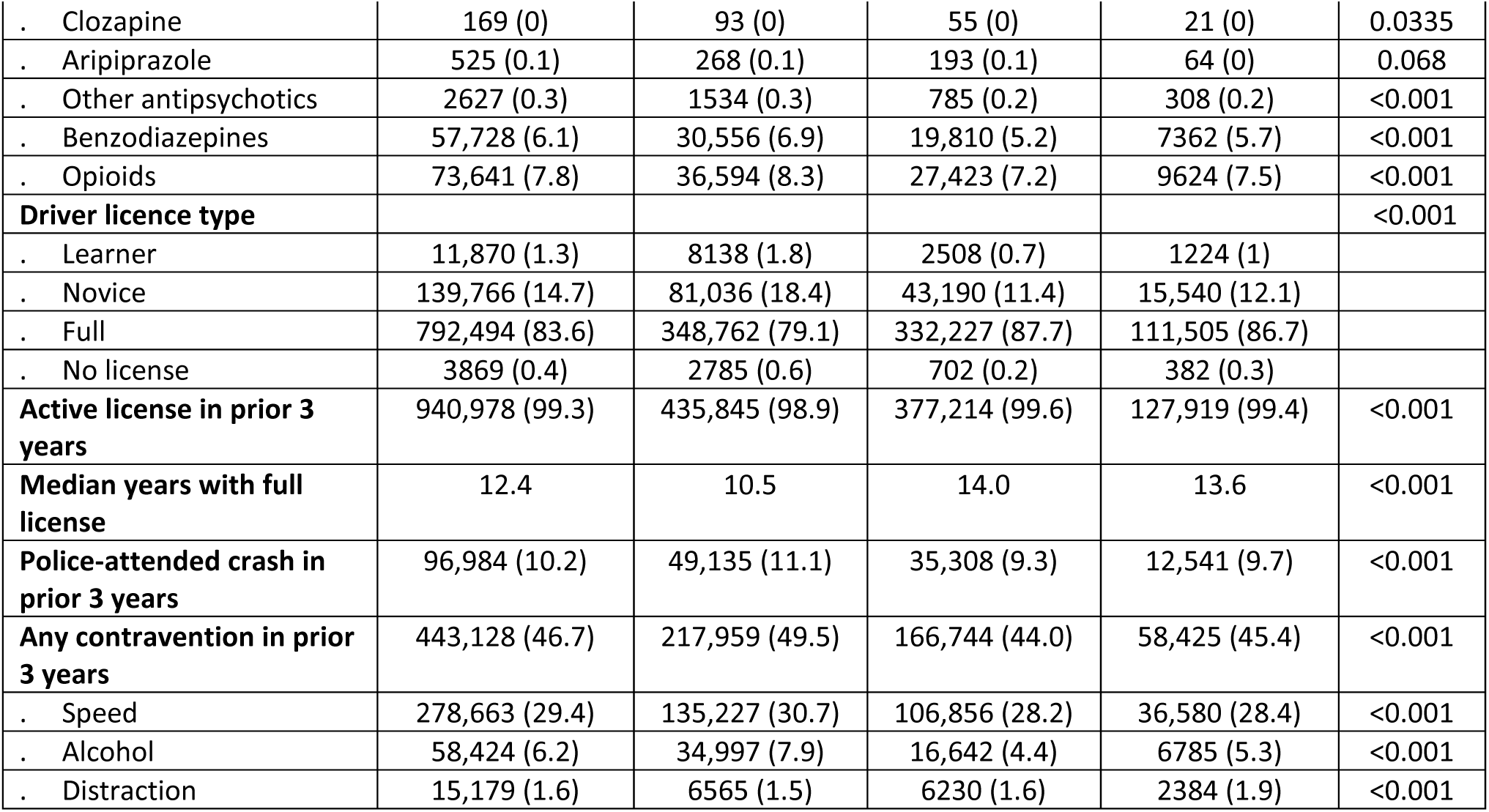
Cohort characteristics for the responsibility analysis. **Legend.** Characteristics of all 947,999 drivers involved in a police-attended crash over the study interval. p-values compare the characteristics between responsible and non-responsible drivers. * Defined as ≥1 hospitalization or ≥2 physician visits for the condition over the prior 3 years.

Drivers deemed responsible for their crash were more likely to have established risk factors for crash that do not influence responsibility scores, supporting the validity of responsibility analysis (e.g., more likely to be younger, male, and a learner or novice license holder; **Table 2**). Police were also much more likely to report suspected impairment by alcohol or drugs at the index crash (unadjusted prevalence, 11.0% vs 2.2% among non-responsible drivers; uOR, 5.58; 95%CI, 5.45-5.71; p<0.001; **eTable 6**).

In total, 178 of 235 drivers with a recent hospitalization for acute psychosis and 440,543 of 819,113 drivers without recent psychosis were deemed responsible for their crash, corresponding to a significant association between recent acute psychosis and crash responsibility (75.7% vs 53.8% deemed responsible; aOR, 2.38; 95%CI, 1.75-3.24; p<0.001; eTable 7).

Results from subgroup and sensitivity analyses were generally consistent with the main results **(eTable 8**, **eFigure 3)**. We observed stronger associations between crash responsibility and recent hospitalization for psychosis for crashes resulting in injury or fatality (aOR, 2.89; 95%CI, 1.85-4.52; p<0.001), when psychosis was attributed to schizophrenia (aOR, 6.20; 95%CI, 2.16-17.77; p<0.001), and when exposure lookback windows were shorter (e.g., with a 2-week exposure lookback from crash, aOR, 3.12; 95%CI, 1.95-4.99; p<0.001).

**Figure 3.**
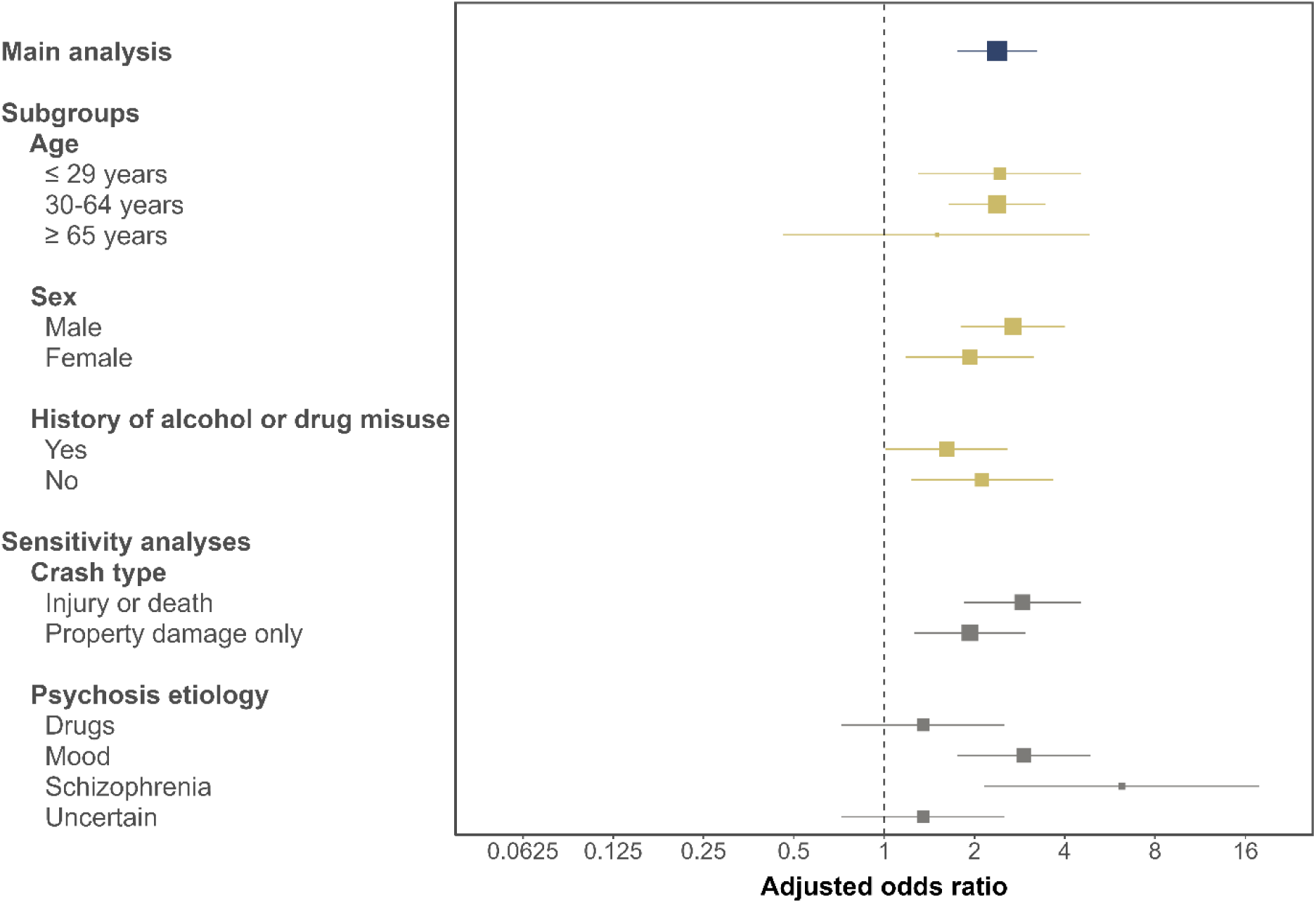
Forest plot of main and subgroup analyses for the responsibility study. **Legend.** Adjusted odds ratios evaluating the association between psychosis and crash responsibility for key subgroup and sensitivity analyses. X-axis depicts the adjusted odds ratio on a log scale; squares, the point estimate, with size reflecting the inverse of the standard error; horizontal lines, the 95% confidence interval. The results of subgroup and sensitivity analyses are generally consistent with results of the main responsibility analysis.

Drivers with psychosis exhibited a lower absolute annual crash risk but a higher responsible crash rate relative to controls, strongly suggesting they have reduced road exposure relative to controls and highlighting a key advantage of responsibility analysis (**eTable 9**).

## Discussion

In a population-based cohort of drivers involved in police-attended crashes over a 17-year study interval, we found an association between recent hospitalization for acute psychosis and subsequent motor vehicle crash. A self-matched case-crossover study of 9842 crash-involved drivers found that a recent hospital stay for acute psychosis was temporally associated with a 1.3-fold increase in the odds of crash. A responsibility analysis examining 819,348 crash-involved drivers found those with a recent hospital stay for acute psychosis had a 2.4-fold increase in the odds of being deemed responsible for their crash. The consistent association across multiple study designs supports the notion that acute psychosis causes a transient increase in crash risk that has not returned to baseline by the time of hospital discharge.^29^ Notably, most studies indicate that antipsychotic medications have a neutral or beneficial effect on crash risk among individuals with psychotic conditions, suggesting it is the psychosis itself that causes the transient increase in crash risk.^29,30,31^ Taken together, these findings suggest that efforts to reduce traffic injury after an episode of acute psychosis are warranted.

Two additional aspects of our findings merit particular consideration. First, hospitalization for psychosis was rare: only 235 of 819,348 drivers with determinate responsibility (0.03%) had a hospitalization for acute psychosis ending in the 6 weeks prior to crash. While steps to reduce traffic injury after an episode of psychosis are warranted, a meaningful reduction in the overall burden of traffic injury among individuals with psychotic disorders will require a broader suite of interventions that target more common crash risk factors such as speeding, failure to use seatbelts, alcohol-imparied driving and distracted driving.^32^ Second, acute psychosis appears to increase crash risk, but context is important. A recent ticket for speeding or distracted driving and recent psychosis were both associated with a similar increase in the odds of subsequent crash (aOR 1.3 vs 1.3); age ≥65 years, possession of a learner license, and recent psychosis all had similar strengths of association with driver responsibility for subsequent crash (aORs 2.0, 2.4, and 2.4, respectively). It would generally be considered disproportionate and unfair for a 65^th^ birthday to prompt an automatic license suspension; similar considerations for fairness and proportionality should apply to individuals recovering from an episode of psychosis, in whom severe restrictions may unnecessarily intensify social and occupational marginalization.^33^ For context, non-zero blood alcohol concentrations <0.05% are thought to double crash risk but are not subject to any penalty in many jurisdictions.^34^

Our study has many strengths. We focused on road safety in the clinically-relevant group of individuals who continue to drive after an episode of psychosis. We used population-based data for a large cohort of crash-involved drivers, yielding highly generalizable results. Use of empirical crash data minimized outcome misascertainment, avoiding biases arising from crash self-reporting and circumventing concerns about the uncertain applicability of driving simulators to real-world road safety. We used population-wide hospital records to identify psychosis hospitalizations, limiting exposure misascertainment. The case-crossover analysis perfectly controlled for relatively fixed individual characteristics (such as driving experience and routine driving behaviours) and adjusted for important time-varying confounders (such as problematic alcohol and drug use). The responsibility analysis fully accounted for road exposure, thus mitigating a major bias of existing studies on driving safety among individuals with psychotic disorders. Analysis of alternate exposure intervals strengthened causal inference, and multiple sensitivity analyses suggested results were robust to changes in study design.

Our study has limitations. Analyses were restricted to drivers involved in a police-attended crash. Results may not apply to individuals who never drive or to those involved in less serious crashes. We identified psychosis requiring hospitalization using comprehensive population-based hospital records, but we could not identify less severe psychosis that was treated as an outpatient or psychosis that did not receive medical attention.^1^ It is possible that psychosis hospitalization in the control interval influences the subsequent likelihood of psychosis hospitalization in the pre-crash interval. We lacked granular data on road exposure. The case-crossover study might find a spurious positive association if individuals drove greater distances in the first few weeks after acute psychosis, but we think it is more likely that psychosis reduces road exposure and that our case-crossover results instead underestimate the association between acute psychosis and crash risk.^5,6,7,8,9,12,13^ We lacked granular clinical data on alcohol and drug use, mental health history, and socioeconomic status. Results may be subject to residual confounding. The specific driving advice provided to each study subject and the degree of adherence to recommendations remains unknown. We focused on private (i.e., non-commercial) drivers and our findings do not apply to professional drivers who have vastly greater road exposure and more severe crash consequences if driving large vehicles or those with many passengers.

Using case-crossover and responsibility analyses to examine all eligible police-attended crashes over a 17-year study interval, we found the 6-week interval following a hospital stay for acute psychosis is associated with increased odds of subsequent crash and increased likelihood of the driver being deemed responsible for the crash. Clinicians should advise all patients recovering from an episode of acute psychosis to wear a seatbelt, leave ample headway, adhere to posted speed limits, avoid distracted driving, and avoid alcohol- and drug-impaired driving.

Clinicians should also thoughtfully consider driving safety when discharging patients from a psychiatric hospitalization, advising patients judged to be at higher risk of crash to limit or temporarily cease driving during their convalescence.

## Data Availability

Access to data provided by the Data Stewards is subject to approval but can be requested for research projects through the Data Stewards or their designated service providers.

