## Supplementary Appendix for "Acute psychosis and the risk of motor vehicle crash"

### eMethods 1: Additional methods

#### Data sources

We accessed health and driver data through Population Data BC, a university-based repository for individual-level longitudinal administrative health data on all BC residents.[1] Probabilistic linkage between driver license number and personal health number (PHN) was based on name, sex, and birthdate and achieved linkage rates ≥95%. All BC residents who held a driver license during the study period were eligible for linkage. Population Data BC deidentified the final dataset before release to investigators through the Secure Research Environment. Our specific data sources included:

| Source of data | Type of data |
|---|---|
| **Health data** | |
| Consolidation File | Registration with BC's provincial health insurance plan |
| Discharge Abstract Database (DAD) | Hospitalization records |
| Medical Service Plan (MSP) | Physician fee-for-service payment claims data |
| PharmaNet | Outpatient prescription fill data |
| Income Band database | Average household income by neighbourhood |
| Vital Statistics database | Date and cause of death |
| **Driving data** | |
| BC Traffic Accident System (TAS) | Police-reported data for all police-attended crashes |
| Insurance Corporation of British Columbia (ICBC) files including the Driver Experience table, the Exam table, the Contraventions table | Driver license data, contraventions data |

- **Discharge Abstract Database**: Canadian Institute for Health Information [creator] (2017): Discharge Abstract Database (Hospital Separations). Population Data BC [publisher]. Data Extract. MOH (2017). http://www.popdata.bc.ca/data.
- **PharmaNet**: BC Ministry of Health [creator] (2018): PharmaNet. V2. BC Ministry of Health [publisher]. Data Extract. Data Stewardship Committee (2018). http://www.popdata.bc.ca/data
- **Medical Services Plan**: British Columbia Ministry of Health [creator] (2017): Medical Services Plan (MSP) Payment Information File. Population Data BC [publisher]. Data Extract. MOH (2017). http://www.popdata.bc.ca/data
- **Consolidation File**: British Columbia Ministry of Health [creator] (2017): Consolidation File (MSP Registration & Premium Billing). Population Data BC [publisher]. Data Extract. MOH (2017). http://www.popdata.bc.ca/data
- **BC Vital Statistics Agency** [creator] (2017): Vital Statistics Deaths. V2. Population Data BC [publisher]. Data Extract BC Vital Statistics Agency (2017). http://www.popdata.bc.ca/data
- **Statistics Canada**, Small Area and Administrative Data Division [creator] (2017): Income Band data file. Population Data BC [publisher]. Data Extract. Statistic Canada (2017). http://www.popdata.bc.ca/data
- **Income Band**: Statistics Canada [creator]: Statistics Canada Income Band Data. Catalogue Number: 13C0016. V2. Population Data BC [publisher]. Data Extract. Population Data BC (2017). http://www.popdata.bc.ca/data
- **Driver data**: Insurance Corporation of British Columbia [creator] (2017): Driver Experience, Contraventions, and Exam tables and the Traffic Accident System. Insurance Corporation of British Columbia [publisher]. Data Extract. ICBC (2017).

All inferences, opinions and conclusions drawn in this manuscript are those of the authors and do not reflect the opinions or policies of the Data Stewards. Access to data provided by the Data Stewards is subject to approval, but can be requested for research projects through the Data Stewards or their designated service providers. Further information regarding these data sets can be found in the PopData project webpage: https://my.popdata.bc.ca/project_listings/13-039/collection_approval_dates

**Missing data**

Study datasets are highly complete for all critical study variables, as described in prior manuscripts[2]:

**a)** Exposure variable (psychosis) was never missing as it was defined by a hospital visit with validated diagnostic codes corresponding to a discharge diagnosis of acute psychosis.[3] When no hospitalization occurred in the exposure lookback period, or when hospitalizations in the exposure lookback period did not have a discharge diagnosis of acute psychosis, the exposure was deemed to be absent. Thus there were no missing data for exposure.

**b)** Outcome (crash in the case-crossover study; crash responsibility in the responsibility study) is almost never missing. Police are required to attend all fatal crashes and almost all serious crashes in BC. Attending police officers report factors that contribute to the crash (e.g. animal on road, speeding, defective brakes). The absence of these factors on the report indicates the officer believed these factors did not contribute to the crash (and thus do not exonerate the driver of responsibility). Less than 2% of TAS police reports are excluded because they are missing data in ≥3 scoring categories.

**c)** Residential variables in the case-crossover analysis (residential neighbourhood household income quintile, residential urbanicity, residential region) had missingness within the acceptable range (<5%). Missing residential variables were imputed using forward/backward filling if present in other pre-crash or control intervals. We think this approach is acceptable as these variables remained static between crash and control dates for most of the cohort. If residential data were missing for all intervals for a driver-crash pair, they were dropped from the case-crossover analysis.

**d)** Breath alcohol level is usually not reported because the attending officer may not obtain a breath alcohol level if they do not suspect impairment. There is evidence of impairment (positive breath alcohol level, police suspicion of impairment by alcohol or drugs) in 6.6% of crash-involved drivers. All other drivers were assumed to have no impairment. Thus there were no missing data for impairment.

**e)** Missingness in other key variables (e.g. age, sex, number of prior hospitalizations, license type, years of driving experience) were within acceptable range (<5%) and did not necessitate the use of multiple imputation.[4,5]

**Independence of events**

Drivers involved in multiple police-attended crashes over the study interval could contribute more than one eligible crash to the analysis. We treated each unique driver-crash combination as an independent observation because police complete crash reports for involved drivers without any information from prior crash reports (making crash responsibility independent of the driver's responsibility for prior crashes), and because each driver involved in a crash is scored independently, with no requirement that one driver be deemed responsible and the others to be deemed non-responsible for the crash.

**A note on responsibility**

In Analysis 2 of this study, the concept of 'crash responsibility' is independent of legal or insurance-based responsibility for the crash. It is not related to "Not Criminally Responsible on Account of Mental Disorder" (NCR), a verdict that can be passed in Canadian courts indicating an individual accused of a crime was "suffering from a mental disorder that rendered the person incapable of appreciating the nature and quality of the act or omission or of knowing that it was wrong".[6]

### eTable 1: ICD-10-CA Diagnostic codes to identify hospitalizations for acute psychosis

| Purpose | Description | ICD-10-CA codes |
|---|---|---|
| Non-schizophrenia psychosis | Mental and behavioural disorders due to use of alcohol – psychotic disorder, residual and late-onset psychotic | F105, F107 |
| | Mental and behavioural disorders due to use of drugs (opioids, cannabinoids, sedatives, cocaine, stimulants, hallucinogens, tobacco, volatile solvents, multi drug use) – psychotic disorder, residual and late-onset psychotic | F115, F117, F125, F127, F135, F137, F145, F147, F155, F157, F165, F167, F175, F177, F185, F187, F195, F197 |
| | Delusional disorder | F220, F228, F229 |
| | Acute polymorphic psychotic disorder | F230, F231 |
| | Acute schizophrenia-like psychotic disorder | F232 |
| | Other acute predominantly delusional and transient psychotic disorders | F233, F238, F239 |
| | Induced delusional disorder | F24 |
| | Other nonorganic psychotic disorders | F28 |
| | Unspecified nonorganic psychosis | F29 |
| | Mood disorders (e.g. mania or depressive episode with psychotic symptoms, bipolar affective) | F302, F312, F315, F232, F333 |
| | Hallucinations (auditory, visual, other, unspecified) | R440, R441, R442, R443, R448 |
| Schizophrenia | Schizophrenia | F20 |
| | Paranoid schizophrenia | F200 |
| | Disorganized schizophrenia | F201 |
| | Catatonic schizophrenia | F202 |
| | Undifferentiated schizophrenia | F203 |
| | Post-schizophrenic depression | F204 |
| | Residual schizophrenia | F205 |
| | Simple schizophrenia | F206 |
| | Other schizophrenia (schizophreniform disorder, other) | F208 |
| | Schizophrenia, unspecified | F209 |
| | Schizoaffective disorders (bipolar type, depressive type, other, unspecified) | F250, F251, F252, F258, F259 |

**Legend**. Asterisk (*) indicates inclusion of the following subtypes: unspecified, subchronic, chronic, subchronic with acute exacerbation, chronic with acute exacerbation, in remission. ICD = International Statistical Classification of Diseases and Related Health Problems.[7] Hospitalizations with a most responsible diagnosis of delirium (ICD-10-CA codes starting with F05) were not considered an exposure because this diagnosis is distinct from psychosis.

### eFigure 1: Study schematics of case-crossover and responsibility analyses study designs

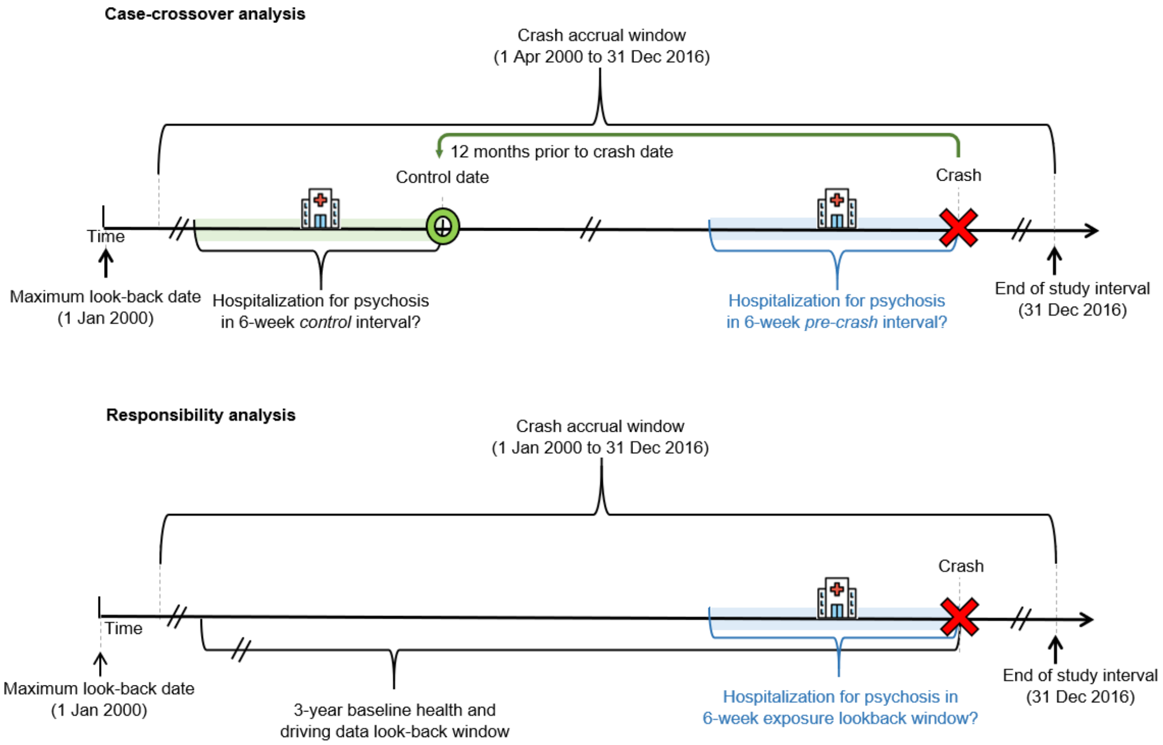

**Legend**: The top panel illustrates the case-crossover study design; the bottom panel illustrates the responsibility analysis study design. Horizontal arrow depicts passage of time from left to right. Anchored on the index crash date (outcome; red X), we used a 6-week exposure lookback window for recent psychosis. We also used a 90-day (in case-crossover) and 3-year (in responsibility analysis) covariate lookback window to identify baseline medical and crash history.

### eFigure 2: Conceptual model of psychosis and crash

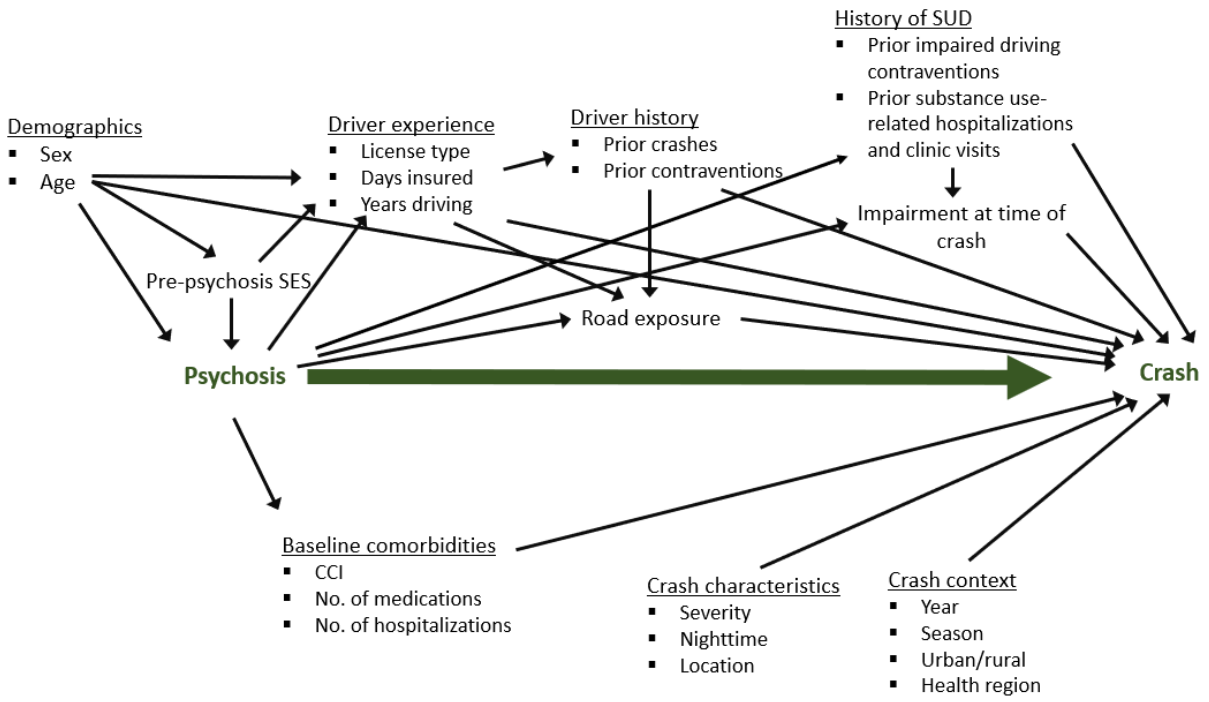

**Legend.** Directed acyclic graph (DAG) depicting causal relationships between psychosis and crash, and potential confounders. SES = socioeconomic status; SUD = substance use disorder; CCI = Charlson comorbidity index. Acute psychosis potentially changes road exposure (hours or miles of driving per week), which in turn influences crash risk per unit time; road exposure can therefore be seen as a mediator of the association of interest. A recent history of 'impairment-related traffic contraventions' (a proxy for impaired driving) can be seen as a mediator of one pathway between psychosis and crash (as illustrated here). However, we choose to adjust for these contraventions " because we wanted to isolate the impact of psychosis itself on crash risk. If the association between psychosis hospitalization and crash risk was entirely due to an increase in impaired driving, this may not motivate any changes in clinical practice or policy because impaired driving is already illegal. In the case-crossover study, unadjusted effect estimates (which still control for fixed confounding factors through self-matching) are similar to adjusted effect estimate, suggesting these potential mediating effects are likely small (eTable 5).

### eTable 2: Variable definitions

| Variables | Description | CCO | RA |
|---|---|---|---|
| **Exposure** | | | |
| Psychosis | Discharge from the hospital for acute psychosis of any etiology in the 6 weeks prior to crash (including psychosis attributed to schizophrenia, substance use, brief psychotic disorder, depressive disorder or bipolar disorder. Obtained via ICD-10-CA codes in the 'Most Responsible Diagnosis' field of DAD; Appendix eTable 1). Binary (yes/no) | x | x |
| **Outcomes** | | | |
| Crash as driver (case-crossover analysis) | Involvement in a police-attended crash as a driver in the crash accrual window. Binary (0=no crash, 1=crash) | x | |
| Crash responsibility (responsibility analysis) | Dichotomized as 'responsible' (score ≤13) or 'non-responsible' (score ≥16). Indeterminate scores (14-15) were excluded from further analysis. Binary (0=non-responsible, 1=responsible) | | x |
| **Confounders** | | | |
| **Demographic** | | | |
| Age | Categorical (<21; 21-44, ref; 45-64; >64 years) | | x |
| Sex | Male; Female (ref); Unknown | | x |
| Residential neighbourhood household income quintile | Neighbourhood income quintile (1 = lowest, 5 = highest) generated by PopDataBC using census data and postal codes (ordinal) | x | x |
| Residential urbanicity | Urban (ref); rural and remote according population density; small town, missing | x | x |
| Residential region | BC's 5 health authority regions: Vancouver Coastal (ref); Vancouver Island; Fraser; Interior; Northern | | x |
| **Health** | | | |
| Number of active medications | Total number of different classes of medications with prescriptions that cover index date, based on fill date and days supplied. Continuous variable. | x | |
| Hospitalizations in lookback period | Total number of hospitalizations in the lookback. Continuous variable. | x | |
| Hospitalization or clinic visits for alcohol misuse in the lookback period | Number of hospitalizations or outpatient physician visits for alcohol misuse disorder in the lookback period. Continuous variable. | x | |
| Hospitalization or clinic visits for non-alcohol substance misuse in the lookback period | Number of hospitalizations or outpatient physician visits for non-alcohol substance use disorder in the lookback period. Continuous variable. | x | |
| Hospitalization or clinic visits for SUD in the lookback period | Number of hospitalizations or outpatient physician visits for substance use disorder in the lookback period. Continuous variable. | | x |
| CCI in the 3y lookback period | Dichotomized variable for Charlson Comorbidity Index ≥2 (yes/no) | | x |
| **Road exposure** | | | |
| License type | Full license (ref), Novice license; Learner license; None | | x |
| Number of years with a full license | Zero if Novice or Learner license types. Continuous variable. | | x |
| **Driving experience** | | | |
| Number of police-attended crashes in 3y lookback period | Continuous variable. | | x |

| | | | |
|---|---|---|---|
| Number of non-impaired contraventions in lookback period | Traffic violations include speeding and distracted driving (but excluding impairment by alcohol or drugs). Continuous variable. | x | x |
| Number of impaired driving events in the lookback period | Traffic violations indicating impairment by alcohol or drugs. Continuous variable. | x | x |
| **Crash characteristics** | | | |
| Crash severity | Property damage only (ref); Injury; Fatality | | x |
| Nighttime crash | Yes if crash occurred between 9pm and 6am. | | x |
| Crash location type | City street (ref); Highway; Rural roads | | x |
| Crash year | Calendar year of index crash | | x |
| Crash season | Season in which index crash occurred: Dec- Feb (Winter, ref); Mar- May (Spring); Jun-Aug (Summer); Sep-Nov (Fall) | | x |
| Impairment by alcohol or drugs | Any documented impairment by drugs and alcohol (police suspicion of impairment, impairment-related contraventions, positive breath alcohol). (yes/no) | | x |

**Legend**. List of exposure, outcomes and candidate confounders included in this study for case-crossover analysis (CCO) and responsibility analysis (RA). Covariates were identified based on expert knowledge, literature review and our direct experience with these data. Variables were selected for inclusion in regression models separately for each study design.

DAD, Discharge Abstract Database; ref, referent variable.

### eTable 3. Crash characteristics for the case-crossover analysis

| Characteristics | All index crashes Count (%) |
|---|---|
| **Crash type** | |
| **Crash severity** | |
| Fatality | 60 (0.6) |
| Injury | 4657 (47.1) |
| Property damage only | 5169 (52.3) |
| **Vehicles involved** | |
| Single-vehicle crash | 2709 (27.4) |
| Multi-vehicle crash | 6045 (61.1) |
| Unknown | 1132 (11.5) |
| **Crash timing** | |
| **Crash year** | |
| 2000-2004 | 3050 (30.9) |
| 2005-2010 | 3965 (40.1) |
| 2011-2016 | 2871 (29) |
| **Crash season** | |
| Spring (Mar-May) | 2447 (24.8) |
| Summer (Jun-Aug) | 2366 (23.9) |
| Fall (Sep-Nov) | 2482 (25.1) |
| Winter (Dec-Feb) | 2591 (26.2) |
| **Crash day-of-week** | |
| Weekend (Fri-Sun) | 5646 (57.1) |
| Weekday (Mon-Thurs) | 4240 (42.9) |
| **Crash time** | |
| Morning (6:01-12:00) | 2324 (23.5) |
| Afternoon (12:01-18:00) | 4043 (40.9) |
| Evening (18:01-24:00) | 2327 (23.5) |
| Night (0:01 - 6:00) | 995 (10.1) |
| Other/unknown | 197 (2) |
| **Contributing factors** | |
| **Impaired by alcohol or drugs** | |
| Yes | 1355 (13.7) |
| No evidence | 8531 (86.3) |
| **Breath alcohol positive** | |
| Yes | 159 (1.6) |
| No evidence | 9727 (98.4) |
| **Speed** | 1234 (12.5) |
| **Human condition** | 3806 (38.5) |
| Distracted/inattentive | 2246 (22.7) |
| Alcohol | 1034 (10.5) |
| Drugs | 319 (3.2) |
| Illness/fatigue | 337 (3.4) |
| Medications | 123 (1.2) |
| Pre-existing physical disability | 51 (0.5) |

### eTable 4. Full regression model results for case-crossover analysis

| Model covariates | Adjusted odds ratio (95%CI) | p-value |
|---|---|---|
| **Exposure: Recent hospital stay for psychosis** | **1.32 (1.05, 1.66)** | **0.018** |
| Residential neighbourhood household income quintile (1 lowest, 5 highest) | 0.96 (0.92, 1.00) | 0.043 |
| Residential urbanicity: small town (ref: urban) | 1.03 (0.83, 1.28) | 0.789 |
| Residential urbanicity: rural (ref: urban) | 1.06 (0.85, 1.31) | 0.621 |
| Number of impaired driving events in the 90 days prior to index date | 2.11 (1.71, 2.59) | <0.001 |
| Number of non-impaired contraventions in the 90 days prior to index date | 1.30 (1.22, 1.37) | <0.001 |
| Number of hospital admissions in the 90 days prior to index date | 1.08 (1.00, 1.17) | 0.050 |
| Number of prescriptions active at index date | 1.10 (1.07, 1.12) | <0.001 |
| Number of hospitalization or clinic visits for non-alcohol substance misuse in the 90 days prior to index | 0.75 (0.41, 1.38) | 0.354 |
| Number of hospitalization or clinic visits for alcohol substance misuse in the 90 days prior to index | 1.73 (0.86, 3.47) | 0.122 |

### eTable 5. Subgroup and sensitivity analyses for crash risk (case-crossover analysis)

| Description | Crash-involved drivers | Exposed at crash | Exposed at control interval 1 | Exposed at control interval 2 | Exposed at control interval 3 | Unadjusted odds ratio (95% CI; p-value) | Adjusted odds ratio (95% CI; p-value) |
|---|---|---|---|---|---|---|---|
| **Primary analysis** | 9842 | 199 | 147 | | | 1.40 (1.12, 1.75); 0.004 | 1.32 (1.05, 1.66); 0.019 |
| **Subgroups** | | | | | | | |
| **Age** | | | | | | | |
| ≤44 years old | 6353 | 137 | 105 | | | 1.34 (1.02, 1.74); 0.032 | 1.22 (0.93, 1.60); 0.157 |
| >44 years old | 3489 | 62 | 42 | | | 1.56 (1.02, 2.36); 0.039 | 1.50 (0.98, 2.30); 0.062 |
| **Sex** | | | | | | | |
| Male | 6355 | 132 | 101 | | | 1.35 (1.02, 1.77); 0.033 | 1.30 (0.98, 1.73); 0.066 |
| Female | 3487 | 67 | 46 | | | 1.50 (1.02, 2.22); 0.042 | 1.36 (0.91, 2.02); 0.129 |
| **History of alcohol or other drug misuse** | | | | | | | |
| Yes | 1551 | 58 | 39 | | | 1.58 (1.02, 2.44); 0.041 | 1.56 (0.99, 2.45); 0.054 |
| No | 8291 | 141 | 108 | | | 1.34 (1.03, 1.74); 0.030 | 1.24 (0.95, 1.61); 0.119 |
| **Impaired at time of crash** | | | | | | | |
| Yes | 1349 | 39 | 19 | | | 2.33 (1.27, 4.27); 0.006 | 2.11 (1.13, 3.95); 0.019 |
| No | 8493 | 160 | 128 | | | 1.28 (1.00, 1.63); 0.050 | 1.21 (0.94, 1.54); 0.139 |
| **Sensitivity analysis: Alternate exposure interval lengths** | | | | | | | |
| 2 weeks | 9842 | 93 | 61 | | | 1.53 (1.11, 2.12); 0.010 | 1.48 (1.06, 2.07); 0.021 |
| 4 weeks | 9842 | 146 | 109 | | | 1.37 (1.06, 1.77); 0.017 | 1.30 (1.00, 1.68); 0.052 |
| 6 weeks (primary analysis) | 9842 | 199 | 147 | | | 1.40 (1.12, 1.75); 0.004 | 1.32 (1.05, 1.66); 0.019 |
| 8 weeks | 9842 | 249 | 190 | | | 1.35 (1.11, 1.65); 0.003 | 1.28 (1.04, 1.56); 0.019 |
| 12 weeks | 9842 | 346 | 259 | | | 1.38 (1.17, 1.64); 2e-04 | 1.31 (1.10, 1.57); 0.002 |
| **Sensitivity analyses: Alternate study designs** | | | | | | | |
| 1 control date = t0-1y (primary analysis) | 9842 | 199 | 147 | | | 1.40 (1.12, 1.75); 0.004 | 1.32 (1.05, 1.66); 0.019 |
| 2 control dates = t0-1y, t0-2y (1 and 2 years prior to crash) | 7874 | 165 | 126 | 98 | | 1.53 (1.24, 1.88); <0.0001 | 1.34 (1.08, 1.67); 0.007 |
| 3 control dates = t0-1y, t0-2y, t0-3y (1, 2, and 3 years prior to crash) | 6288 | 137 | 100 | 83 | 84 | 1.59 (1.28, 1.97); <0.0001 | 1.35 (1.08, 1.68); 0.008 |
| **Sensitivity analysis: Alternate outcome** | | | | | | | |
| Only casualty crashes (fatality or injury) | 4695 | 100 | 71 | | | 1.44 (1.05, 1.97); 0.023 | 1.32 (0.96, 1.82); 0.087 |
| Only property damage crashes | 5147 | 99 | 76 | | | 1.35 (0.98, 1.87); 0.064 | 1.32 (0.95, 1.83); 0.097 |

**Legend.** Subgroup analyses explored whether the observed association varied across clinically relevant strata. Sensitivity analyses explored alternate exposure interval lengths (because the typical duration of elevated crash risk after psychosis hospitalization was unknown), alternate numbers of control dates (because results generated using multiple control dates might signal bias or insufficient precision in the main analysis), and alternate outcomes (because crash severity is meaningful to drivers, patients, clinicians and policymakers). In the sensitivity analysis that explored alternate study designs, the number of crash-involved drivers is smaller in analyses with a greater number of matched control dates because we excluded drivers that did not have an active license during all control periods.

### eTable 6. Selected crash characteristics among responsible and non-responsible drivers

| Characteristics | Responsible drivers N=440,721 Count (%) | Non-responsible drivers N=378,627 Count (%) | p-value |
|---|---|---|---|
| **Basic crash characteristics** | | | |
| **Crash severity** | | | <0.001 |
| Injury or fatality | 205,048 (46.5) | 182,739 (48.3) | |
| Property damage only | 235,673 (53.5) | 195,888 (51.7) | |
| **Vehicle damage severity** | | | <0.001 |
| Light | 94,246 (21.4) | 109,123 (28.8) | |
| Moderate | 123,031 (27.9) | 121,142 (32.0) | |
| Severe | 114,069 (25.9) | 83,210 (22.0) | |
| Demolished | 50,187 (11.4) | 25,758 (6.8) | |
| None | 27,002 (6.1) | 14,353 (3.8) | |
| Other/unknown | 32,186 (7.3) | 25,041 (6.6) | |
| **Number of vehicles involved** | | | <0.001 |
| 1 | 127,672 (29.0) | 64,062 (16.9) | |
| 2 | 269,529 (61.2) | 238,092 (62.9) | |
| 3+ | 43,520 (9.9) | 76,473 (20.2) | |
| **Crash year** | | | <0.001 |
| 2000-2004 | 157,230 (35.7) | 121,873 (32.2) | |
| 2005-2010 | 161,233 (36.6) | 139,623 (36.9) | |
| 2011-2016 | 122,258 (27.7) | 117,131 (30.9) | |
| **Crash season** | | | <0.001 |
| Winter (Dec-Feb) | 106,769 (24.2) | 108,770 (28.7) | |
| Spring (Mar-May) | 104,489 (23.7) | 82,571 (21.8) | |
| Summer (Jun-Aug) | 115,988 (26.3) | 84,204 (22.2) | |
| Fall (Sep-Nov) | 113,475 (25.7) | 103,082 (27.2) | |
| **Day of week** | | | <0.001 |
| Weekday (Mon-Thurs) | 249,091 (56.5) | 221,681 (58.5) | |
| Weekend (Fri-Sun) | 191,630 (43.5) | 156,946 (41.5) | |
| **Time of day** | | | <0.001 |
| Morning (6:01-12:00) | 110,972 (25.2) | 100,331 (26.5) | |
| Afternoon (12:01-18:00) | 187,192 (42.5) | 164,578 (43.5) | |
| Evening (18:01-24:00) | 94,985 (21.6) | 84,596 (22.3) | |
| Night (0:01 - 6:00) | 38,356 (8.7) | 21,709 (5.7) | |
| Other/unknown | 9216 (2.1) | 7413 (2.0) | |
| **Road location** | | | <0.001 |
| City street | 304,254 (69) | 251,213 (66.3) | |
| Provincial highway | 115,554 (26.2) | 113,533 (30.0) | |
| Rural road | 20,913 (4.7) | 13,881 (3.7) | |
| **Crash location** | | | <0.001 |
| At intersection | 196,713 (44.6) | 198,940 (52.5) | |
| Between intersection | 171,060 (38.8) | 144,523 (38.2) | |
| Parking lot | 31,691 (7.2) | 6603 (1.7) | |
| Other/unknown | 41,257 (9.4) | 28,561 (7.5) | |
| **Roadside hazard/design listed as contributory factor** | 2150 (0.5) | 5480 (1.4) | <0.001 |

### eTable 6. Selected crash characteristics among responsible and non-responsible drivers (continued)

| Characteristics | Responsible drivers N=440,721 Count (%) | Non-responsible drivers N=378,627 Count (%) | p-value |
|---|---|---|---|
| **Driving conditions** | | | |
| **Road condition** | | | <0.001 |
| Dry | 288,763 (65.5) | 207,548 (54.8) | |
| Wet | 113,964 (25.9) | 118,618 (31.3) | |
| Snow/slush/ice/mud | 33,749 (7.7) | 50,160 (13.2) | |
| Other/unknown | 4245 (1.0) | 2301 (0.6) | |
| **Road surface** | | | <0.001 |
| Asphalt/concrete | 427,702 (97.0) | 370,414 (97.8) | |
| Stone/gravel/earth/wood | 11,601 (2.6) | 7234 (1.9) | |
| Other/unknown | 1418 (0.3) | 979 (0.3) | |
| **Weather** | | | <0.001 |
| Clear/cloudy | 353,148 (80.1) | 275,119 (72.7) | |
| Rain/strong wind | 65,442 (14.8) | 74,175 (19.6) | |
| Fog/smoke/smog | 3035 (0.7) | 4242 (1.1) | |
| Snow/sleet/hail | 14,303 (3.2) | 22,566 (6.0) | |
| Other/unknown | 4793 (1.1) | 2525 (0.7) | |
| **Lighting** | | | <0.001 |
| Daylight | 295,606 (67.1) | 248,815 (65.7) | |
| Dusk/dawn | 26,223 (6.0) | 24,804 (6.6) | |
| Dark with full illumination | 33,969 (7.7) | 27,194 (7.2) | |
| Dark with no/some illumination | 81,733 (18.5) | 75,995 (20.1) | |
| Other/unknown | 3190 (0.7) | 1819 (0.5) | |
| **Weather/visibility listed as contributory factor** | 27,280 (6.2) | 29,361 (7.8) | <0.001 |
| **Vehicle condition listed as contributory factor** | 2983 (0.7) | 12,269 (3.2) | <0.001 |
| **Unsafe driving actions: Index driver driving safely, obeying road laws** | 75,776 (17.2) | 376,700 (99.5) | <0.001 |
| **Contribution from other parties** | | | <0.001 |
| Yes | 0 (0.0) | 240,941 (63.6) | |
| No/index driver driving unsafely | 440,721 (100.0) | 137,686 (36.4) | |

### eTable 6. Selected crash characteristics among responsible and non-responsible drivers (continued)

| Characteristics | Responsible drivers N=440,721 Count (%) | Non-responsible drivers N=378,627 Count (%) | p-value |
|---|---|---|---|
| **Task involved** | | | |
| **Avoidance maneuver listed as contributory factor** | 3152 (0.7) | 8579 (2.3) | <0.001 |
| **Pre-collision action** | | | <0.001 |
| Straight | 238,055 (54.0) | 247,112 (65.3) | |
| Backing | 26,904 (6.1) | 2413 (0.6) | |
| Turning | 101,716 (23.1) | 38,673 (10.2) | |
| Changing lanes/merging | 9446 (2.1) | 2450 (0.6) | |
| Loss of control | 10,350 (2.3) | 4385 (1.2) | |
| Stopped/parked | 8673 (2.0) | 58,591 (15.5) | |
| Other/unknown | 45,577 (10.3) | 25,003 (6.6) | |
| **Other crash characteristics** | | | |
| **Human condition as contributory factor** | | | |
| Alcohol | 43,684 (9.9) | 6766 (1.8) | <0.001 |
| Medications | 1247 (0.3) | 197 (0.1) | <0.001 |
| Drugs | 4623 (1.0) | 738 (0.2) | <0.001 |
| Illness/fatigue | 7815 (1.8) | 1350 (0.4) | <0.001 |
| Distracted/inattentive | 121,598 (27.6) | 33,704 (8.9) | <0.001 |
| Pre-existing physical disability | 1326 (0.3) | 273 (0.1) | <0.001 |
| **Breath alcohol positive** | | | <0.001 |
| Yes | 7097 (1.6) | 1539 (0.4) | |
| No evidence | 433,624 (98.4) | 377,088 (99.6) | |
| **Impaired by alcohol or drugs** | | | <0.001 |
| Yes | 48,681 (11.0) | 8243 (2.2) | |
| No evidence | 392,040 (89.0) | 370,384 (97.8) | |
| **Speed zone** | | | <0.001 |
| < 50 km/h | 26,294 (6.0) | 14,070 (3.7) | |
| 50 km/h | 237,384 (53.9) | 209,707 (55.4) | |
| 60-70 km/h | 58,070 (13.2) | 55,755 (14.7) | |
| 80+ km/h | 70,683 (16.0) | 73,560 (19.4) | |
| Other/unknown | 48,290 (11.0) | 25,535 (6.7) | |

### eTable 7. Full regression model results for responsibility analysis

| Model covariates | Adjusted odds ratio (95%CI) | p-value |
|---|---|---|
| **Exposure: Recent hospital stay for psychosis** | **2.38 (1.75, 3.24)** | **<0.001** |
| Driver age <21 years (ref: 21-44 years) | 1.43 (1.40, 1.45) | <0.001 |
| Driver age 45-64 years (ref: 21-44 years) | 1.02 (1.01, 1.03) | 0.004 |
| Driver age ≥65 years (ref: 21-44 years) | 2.03 (1.99, 2.06) | <0.001 |
| Male sex (ref: female) | 1.04 (1.03, 1.05) | <0.001 |
| Unknown sex (ref: female) | 1.45 (0.95, 2.22) | 0.082 |
| Residential neighbourhood household income quintile (1 lowest, 5 highest) | 0.98 (0.97, 0.98) | <0.001 |
| Residential urbanicity: small town (ref: urban) | 1.02 (1.00, 1.03) | 0.014 |
| Residential urbanicity: rural (ref: urban) | 1.05 (1.04, 1.07) | <0.001 |
| Residential urbanicity: missing (ref: urban) | 1.11 (1.00, 1.24) | 0.051 |
| Fraser health authority (ref: Vancouver Coastal) | 0.96 (0.95, 0.98) | <0.001 |
| Interior health authority (ref: Vancouver Coastal) | 0.96 (0.94, 0.98) | <0.001 |
| Vancouver Island health authority (ref: Vancouver Coastal) | 0.97 (0.95, 0.98) | 0.000 |
| Northern health authority (ref: Vancouver Coastal) | 0.81 (0.79, 0.83) | <0.001 |
| Missing health authority (ref: Vancouver Coastal) | 0.92 (0.83, 1.02) | 0.099 |
| Learner license type (ref: full license) | 2.39 (2.28, 2.51) | <0.001 |
| Novice license type (ref: full license) | 1.44 (1.42, 1.47) | <0.001 |
| No license at time of crash (ref: full license) | 2.77 (2.54, 3.02) | <0.001 |
| Number of years with full license (0 if non-full license at crash) | 0.99 (0.99, 0.99) | <0.001 |
| Number of crashes in the 3 years prior to index crash | 1.09 (1.08, 1.11) | <0.001 |
| Number of non-impairment contraventions in the 3 years prior to index crash | 1.08 (1.08, 1.09) | <0.001 |
| Number of impairment-related contraventions in 3 years prior to index crash | 1.24 (1.22, 1.25) | <0.001 |
| Accident resulted in severe injury (ref: property damage) | 0.94 (0.94, 0.95) | <0.001 |
| Accident resulted in fatality (ref: property damage) | 1.19 (1.13, 1.26) | <0.001 |
| Crash occurred at night | 0.88 (0.87, 0.89) | <0.001 |
| Crash occurred on provincial highway (ref: city streets) | 0.87 (0.86, 0.88) | <0.001 |
| Crash occurred on rural road (ref: city streets) | 1.07 (1.05, 1.10) | <0.001 |
| Any documented impairment by drugs/alcohol at crash | 5.36 (5.23, 5.50) | <0.001 |
| Hospitalizations for substance use disorder in 3 years prior to index crash | 1.02 (1.02, 1.02) | <0.001 |
| Charlson comorbidity index ≥2 in 3 years prior to index crash | 1.02 (1.01, 1.03) | <0.001 |
| Year of the crash | 0.99 (0.99, 0.99) | <0.001 |
| Crash occurred in the spring (Mar-May) (ref: Winter) | 1.26 (1.24, 1.27) | <0.001 |
| Crash occurred in the summer (Jun - Aug) (ref: Winter) | 1.37 (1.35, 1.39) | <0.001 |
| Crash occurred in the fall (Sep - Dec) (ref: Winter) | 1.10 (1.08, 1.11) | <0.001 |

### eTable 8. Subgroup and sensitivity analyses for crash responsibility

| Description | All crash-involved drivers Count (%) | Crash responsible Count (%) | Crash non-responsible Count (%) | Responsible & exposed Count (%) | Non-responsible & exposed Count (%) | Unadjusted odds ratio (95% CI) | Adjusted odds ratio (95% CI) |
|---|---|---|---|---|---|---|---|
| **Primary analysis** | 947,999 (100) | 440,721 (46.5) | 378,627 (39.9) | 178 (0.0) | 57 (0.0) | 2.68 (1.99, 3.62), p<0.001 | 2.38 (1.75, 3.24), p<0.001 |
| **Subgroups** | | | | | | | |
| **Age** | | | | | | | |
| >30 years | 306,799 (32.4) | 164,483 (53.6) | 105,344 (34.3) | 50 (0.0) | 13 (0.0) | 2.46 (1.34, 4.54), p=0.003 | 2.43 (1.30, 4.53), p=0.005 |
| 30-64 | 542,605 (57.2) | 223,524 (41.2) | 240,106 (44.3) | 118 (0.1) | 40 (0.0) | 3.17 (2.22, 4.54), p<0.001 | 2.38 (1.64, 3.45), p<0.001 |
| 65+ | 98,595 (10.4) | 52,714 (53.5) | 33,177 (33.6) | 10 (0.0) | <5 | 1.57 (0.49, 5.02), p=0.44 | 1.50 (0.46, 4.85), p=0.50 |
| **Sex** | | | | | | | |
| Male | 604,119 (63.7) | 287,174 (47.5) | 235,221 (38.9) | 126 (0.0) | 32 (0.0) | 3.23 (2.19, 4.75), p<0.001 | 2.69 (1.80, 4.01), p<0.001 |
| Female | 343,773 (36.3) | 153,481 (44.6) | 143,372 (41.7) | 52 (0.0) | 25 (0.0) | 1.94 (1.21, 3.13), p=0.006 | 1.93 (1.18, 3.15), p=0.008 |
| **History of alcohol or other drug misuse** | | | | | | | |
| Yes | 23,077 (2.4) | 14,067 (61.0) | 6351 (27.5) | 83 (0.6) | 24 (0.4) | 1.56 (0.99, 2.47), p=0.054 | 1.62 (1.01, 2.58), p=0.04 |
| No | 912,450 (96.3) | 420,052 (46.0) | 367,962 (40.3) | 46 (0.0) | 19 (0.0) | 2.12 (1.24, 3.62), p=0.006 | 2.12 (1.23, 3.67), p=0.007 |

### eTable 8. Subgroup and sensitivity analyses for crash responsibility (continued)

| Description | All crash-involved drivers Count (%) | Crash responsible Count (%) | Crash non-responsible Count (%) | Responsible & exposed Count (%) | Non-responsible & exposed Count (%) | Unadjusted odds ratio (95% CI) | Adjusted odds ratio (95% CI) |
|---|---|---|---|---|---|---|---|
| **Sensitivity analyses** | | | | | | | |
| **Subset of outcome definition** | | | | | | | |
| Casualty crashes (fatality or injury) | 450,684 (47.5) | 205,048 (45.5) | 182,739 (40.5) | 94 (0.0) | 26 (0.0) | 3.22 (2.09, 4.97), p<0.001 | 2.89 (1.85, 4.52), p<0.001 |
| Property damage crashes only | 497,315 (52.5) | 235,673 (47.4) | 195,888 (39.4) | 84 (0.0) | 31 (0.0) | 2.25 (1.49, 3.40), p<0.001 | 1.93 (1.26, 2.96), p=0.003 |
| **Subset of exposure: specific etiology of psychosis** | | | | | | | |
| Exposure: psychosis attributed to substance use | 947,999 (100) | 440,721 (46.5) | 378,627 (39.9) | 38 (0.0) | 15 (0.0) | 2.18 (1.20, 3.95), p=0.01 | 1.35 (0.72, 2.52), p=0.35 |
| Exposure: psychosis attributed to mood disorder | 947,999 (100) | 440,721 (46.5) | 378,627 (39.9) | 67 (0.0) | 20 (0.0) | 2.88 (1.75, 4.74), p<0.001 | 2.92 (1.75, 4.88), p<0.001 |
| Exposure: psychosis attributed to schizophrenia or related disorder | 947,999 (100) | 440,721 (46.5) | 378,627 (39.9) | 31 (0.0) | <5 | 6.66 (2.36, 18.78), p<0.001 | 6.20 (2.16, 17.77), p<0.001 |
| Exposure: psychosis with uncertain etiology | 947,999 (100) | 440,721 (46.5) | 378,627 (39.9) | 38 (0.0) | 15 (0.0) | 2.18 (1.20, 3.95), p=0.01 | 1.35 (0.72, 2.52), p=0.35 |
| **Sensitivity analyses - alternate exposure lookback window** | | | | | | | |
| 2 weeks | 947,999 (100) | 440,721 (46.5) | 378,627 (39.9) | 88 (0.0) | 23 (0.0) | 3.29 (2.08, 5.20), p<0.001 | 3.12 (1.95, 4.99), p<0.001 |
| 4 weeks | 947,999 (100) | 440,721 (46.5) | 378,627 (39.9) | 136 (0.0) | 39 (0.0) | 3.00 (2.10, 4.28), p<0.001 | 2.78 (1.93, 4.01), p<0.001 |
| 8 weeks | 947,999 (100) | 440,721 (46.5) | 378,627 (39.9) | 224 (0.1) | 70 (0.0) | 2.75 (2.10, 3.60), p<0.001 | 2.41 (1.83, 3.18), p<0.001 |
| 12 weeks | 947,999 (100) | 440,721 (46.5) | 378,627 (39.9) | 302 (0.1) | 98 (0.0) | 2.65 (2.11, 3.33), p<0.001 | 2.36 (1.86, 2.98), p<0.001 |
| >12 weeks | 947,999 (100) | 440,721 (46.5) | 378,627 (39.9) | 2954 (0.7) | 1497 (0.4) | 1.70 (1.60, 1.81), p<0.001 | 1.70 (1.59, 1.81), p<0.001 |
| Any time prior to crash | 947,999 (100) | 440,721 (46.5) | 378,627 (39.9) | 3256 (0.7) | 1595 (0.4) | 1.76 (1.66, 1.87), p<0.001 | 1.74 (1.64, 1.85), p<0.001 |

**Legend.** Subgroup analyses explored whether the observed association varied across clinically relevant strata. Sensitivity analyses explored crash severity (because this is meaningful to drivers, patients, clinicians and policymakers), the etiology of the acute psychosis (because this is relevant to clinicians), and alternate exposure interval lengths (because the typical duration of elevated crash risk after psychosis hospitalization was unknown).

### eFigure 3. Strength of the association between psychosis and responsibility for crash appears accentuated at shorter exposure lookback intervals

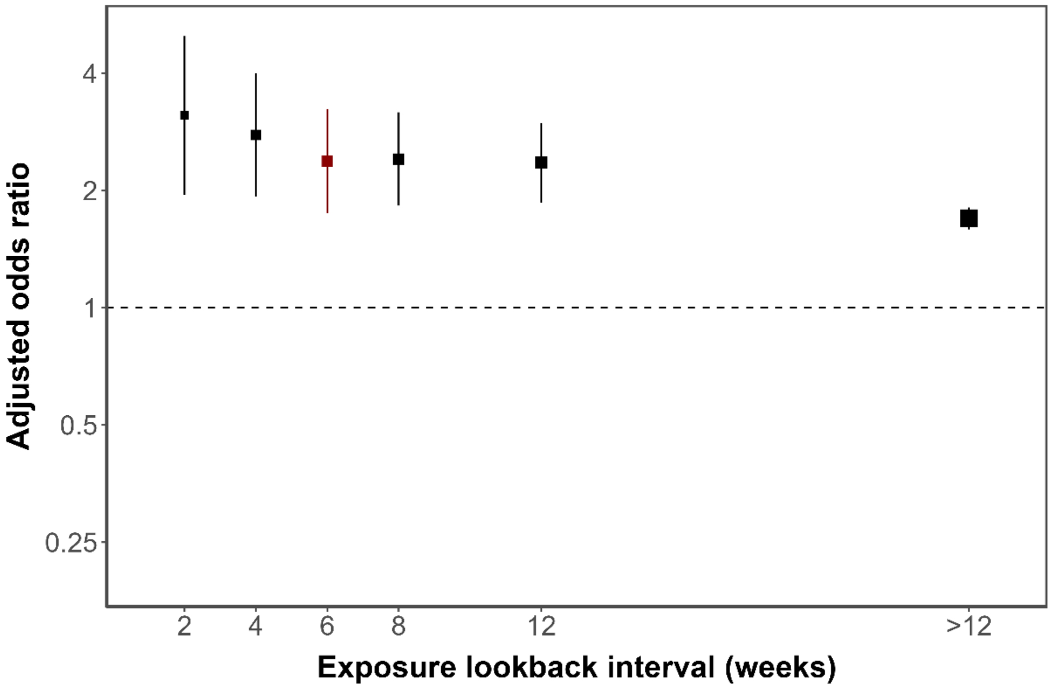

**Legend.** X-axis depicts the exposure lookback interval in weeks; Y-axis depicts the adjusted odds ratio describing the association between driver responsibility for crash and recent hospital stay for acute psychosis; squares depict the point estimate for each sensitivity analysis, aligned with the corresponding exposure lookback; sizes of the squares reflect the inverse of the standard error; vertical lines depict the 95% confidence interval for the adjusted odds ratio. The highest adjusted odds ratio point estimate is observed in the first 2 weeks following discharge, with a gradual reduction over time.

### eTable 9. Comparison of crash rates

| Value | Individuals with psychosis | Controls |
|---|---|---|
| Number of unique drivers | 38,422 | 4,741,298 |
| Number of unique crash-involved drivers | 260 | 757,123 |
| Number of unique driver-crash combination | 264 | 947,735 |
| Number of responsible driver-crash combination | 178 | 440,543 |
| Number of total person-years driving | 498,307 | 60,948,687 |
| Absolute crash rate per driver | 0.007 | 0.200 |
| **Absolute crash rate per driver-year** | **0.001** | **0.016** |
| **Responsible crash rate** | **0.674** | **0.465** |

**Legend**. This analysis first stratified all individuals by the presence of a hospitalization for psychosis at any time during the study interval. Crash rate per driver was calculated by dividing the number of driver-crash combinations by the number of drivers. Crash rate per driver-year is calculated by dividing the number of driver-crash combinations by the number of driver-years of exposure (driver years are calculated by summing the total number of years each driver held an active license in BC during our study interval). Responsible crash rate is calculated by dividing the *number of responsible driver-crash combinations* by the *total number of driver-crash combinations* (note that results presented here include driver-crash combinations with indeterminate crash responsibility in the denominator and consequently and intentionally differ slightly from the responsible crash rates reported in the manuscript text). As expected, absolute crash rates are much higher for controls but responsible crash rates are higher for individuals with prior psychosis, reflecting reduced road exposure but higher crash risk while driving among individuals with psychosis. This finding highlights a major advantage of using responsibility analysis to evaluate whether acute psychosis poses a risk for crash.
